# Bayesian Joint Longitudinal-Survival Modeling of Functional Recovery Trajectories and Time to Independent Community Ambulation Following Robotic Exoskeleton-Assisted Stroke Rehabilitation: A Multi-Centre Cohort Study in Canada

**DOI:** 10.64898/2026.03.12.26348287

**Authors:** Amaevia Lim, Priya Desai

## Abstract

**Background:** Robotic exoskeleton-assisted gait training (RAGT) has emerged as a promising modality for post-stroke rehabilitation. However, the longitudinal trajectory of functional recovery and its association with clinically meaningful milestones such as independent community ambulation remain poorly characterised. Standard analytical approaches that treat longitudinal and survival outcomes separately may yield biased estimates due to informative dropout and the endogenous nature of time-varying functional measures.

**Objective:** To jointly model the longitudinal trajectory of lower-extremity motor function and time to independent community ambulation following RAGT for stroke survivors, while accounting for the mutual dependence between the two processes using a Bayesian joint modelling framework.

**Methods:** A multi-centre retrospective cohort study was conducted across four Canadian rehabilitation hospitals (2019 to 2024). A total of 327 adults with first-ever ischaemic or haemorrhagic stroke who received RAGT using the EksoNR or Indego exoskeleton platforms were included. The primary longitudinal outcome was the Fugl-Meyer Assessment Lower Extremity (FMA-LE) score measured at baseline, weeks 4, 8, 12, 24, and 52. The survival outcome was time to achieving independent community ambulation (Functional Ambulation Category score of 4 or higher sustained for at least two consecutive assessments). A Bayesian shared-parameter joint model was specified, linking a nonlinear mixed-effects longitudinal submodel to a Weibull proportional hazards survival submodel through the current value and slope of the subject-specific FMA-LE trajectory. Estimation was performed using Hamiltonian Monte Carlo sampling with four chains of 5,000 iterations each (2,500 warmup).

**Results:** The median age was 62.4 years (IQR 54.1 to 71.8), 58.1% were male, and 63.0% had ischaemic stroke aetiology. The longitudinal submodel revealed a nonlinear recovery pattern best described by a three-knot restricted cubic spline, with rapid improvement during the first 12 weeks (mean gain 8.7 FMA-LE points, 95% CrI 7.2 to 10.3) followed by a plateau phase. The association parameter linking the current FMA-LE value to the hazard of achieving community ambulation was 0.084 (95% CrI 0.061 to 0.109), indicating that each one-point increase in the subject-specific FMA-LE trajectory was associated with an 8.8% increase in the instantaneous hazard (HR = 1.088, 95% CrI 1.063 to 1.115). The trajectory slope parameter was also significant (0.043, 95% CrI 0.012 to 0.078), suggesting that patients with steeper recovery gradients had additional survival advantages beyond their current functional level. At 52 weeks, 54.7% of participants achieved independent community ambulation. Haemorrhagic stroke (HR = 0.68, 95% CrI 0.49 to 0.93), older age (HR per decade = 0.81, 95% CrI 0.70 to 0.94), and higher baseline NIHSS score (HR per point = 0.94, 95% CrI 0.91 to 0.97) were associated with lower hazards of achieving the ambulation milestone.

**Conclusions:** The Bayesian joint model revealed that both the current functional level and the rate of functional change are independently predictive of achieving community ambulation following RAGT. These findings support individualised rehabilitation planning where treatment intensity may be dynamically adjusted based on the evolving recovery trajectory, and provide further evidence for the clinical value of robotic exoskeleton interventions in stroke rehabilitation.

## 1. INTRODUCTION

Stroke remains a leading cause of long-term disability in Canada and worldwide, with approximately 62,000 new cases annually across Canadian provinces (Public Health Agency of Canada, 2023). Among the constellation of post-stroke impairments, gait dysfunction represents one of the most debilitating sequelae, affecting an estimated 60% to 80% of stroke survivors and substantially limiting participation in community activities (Balami et al., 2011; Jorgensen et al., 1995). The restoration of independent community ambulation is consequently regarded as a primary rehabilitation goal and a key determinant of quality of life and healthcare resource utilisation in this population.

Over the past decade, robotic exoskeleton-assisted gait training (RAGT) has gained increasing traction as an adjunct or alternative to conventional overground gait therapy for stroke survivors. Systematic reviews and meta-analyses have reported moderate to large effect sizes favouring RAGT in improving gait speed, functional ambulation, and lower-extremity motor function (Mehrholz et al., 2020; Molteni et al., 2021). From a health economic perspective, Shankar et al. (2025) demonstrated that robotic exoskeleton therapy achieved acceptable cost-effectiveness thresholds compared with conventional physiotherapy for stroke rehabilitation, with favourable incremental cost-effectiveness ratios under specific clinical scenarios. However, the majority of existing efficacy studies have employed pre-post comparisons or between-group mean differences at fixed timepoints, which do not adequately capture the dynamic, nonlinear nature of functional recovery following RAGT.

A fundamental methodological challenge in stroke rehabilitation research is the interdependence between longitudinal recovery trajectories and the occurrence of clinically meaningful milestones. Standard mixed-effects models for repeated measures assume that dropout is non-informative (i.e., missing at random conditional on observed covariates), whereas survival models for time-to-event outcomes do not incorporate the evolving longitudinal profile as a time-varying predictor. When these two processes are correlated, as is typical in rehabilitation settings where patients who recover faster are more likely to achieve functional milestones earlier and subsequently alter their treatment intensity or monitoring schedule, separate analyses may produce biased and inefficient estimates (Rizopoulos, 2012; Tsiatis and Davidian, 2004).

Joint longitudinal-survival models address this limitation by simultaneously estimating the longitudinal trajectory and the hazard of the time-to-event outcome, linking them through shared random effects or current-value associations (Wulfsohn and Tsiatis, 1997; Henderson et al., 2000). The Bayesian approach to joint modelling offers several additional advantages, including natural uncertainty quantification through posterior credible intervals, the ability to incorporate informative priors from previous studies, and robust estimation even with moderate sample sizes and complex model structures (Ibrahim et al., 2001; Guo and Carlin, 2004).

Despite these methodological strengths, Bayesian joint longitudinal-survival models have rarely been applied in the robotic rehabilitation literature. To our knowledge, no study has examined the association between the evolving functional recovery trajectory following RAGT and the probability of achieving independent community ambulation using this framework. Understanding this association is critical for developing personalised rehabilitation protocols, where treatment intensity and duration may be dynamically tailored based on a patient’s predicted recovery trajectory and milestone attainment probability.

The objectives of this study were threefold: (a) to characterise the longitudinal trajectory of lower-extremity motor function following RAGT using a flexible nonlinear mixed-effects model; (b) to estimate the time-to-event distribution for achieving independent community ambulation; and (c) to quantify the association between the subject-specific recovery trajectory and the hazard of milestone attainment using a Bayesian shared-parameter joint model, adjusting for patient-level and stroke-level covariates.

## 2. METHODS

### 2.1 Study Design and Setting

This was a multi-centre retrospective cohort study conducted across four academic rehabilitation hospitals in Canada: Toronto Rehabilitation Institute (Toronto, Ontario), GF Strong Rehabilitation Centre (Vancouver, British Columbia), Institut de Readaptation Gingras-Lindsay de Montreal (Montreal, Quebec), and Glenrose Rehabilitation Hospital (Edmonton, Alberta). The study period extended from January 2019 to December 2024. The protocol was approved by the Research Ethics Board at each participating site (lead REB: University Health Network, REB No. 2023-0412) and adhered to the Declaration of Helsinki. Given the retrospective design using de-identified clinical data, a waiver of individual consent was granted.

### 2.2 Participants

Eligible participants were adults aged 18 years or older who sustained a first-ever ischaemic or haemorrhagic stroke confirmed by neuroimaging (CT or MRI), were admitted to an inpatient or outpatient rehabilitation programme within 6 months of stroke onset, and received a minimum of 12 sessions of RAGT using either the EksoNR (Ekso Bionics, Richmond, CA) or Indego (Parker Hannifin, Cleveland, OH) robotic exoskeleton platforms. Exclusion criteria included prior stroke, pre-existing neurological conditions affecting gait (e.g., Parkinson disease, multiple sclerosis), bilateral lower-extremity amputation, severe cardiopulmonary contraindications to exoskeleton use, and fewer than two post-baseline FMA-LE assessments. From an initial pool of 489 patients identified through institutional RAGT databases, 327 met all inclusion criteria and comprised the analytical sample.

### 2.3 Outcome Measures

The primary longitudinal outcome was the Fugl-Meyer Assessment Lower Extremity (FMA-LE) subscale, a 34-point ordinal measure of lower-extremity motor impairment with established reliability and validity in stroke populations (Fugl-Meyer et al., 1975; Sullivan et al., 2011). FMA-LE assessments were extracted from clinical records at baseline (prior to first RAGT session) and at standardised assessment timepoints of 4, 8, 12, 24, and 52 weeks post-RAGT initiation. The primary survival outcome was time to achieving independent community ambulation, operationally defined as attaining a Functional Ambulation Category (FAC) score of 4 or higher (indicating independent ambulation on level surfaces) sustained for at least two consecutive clinic assessments separated by a minimum of 4 weeks (Mehrholz et al., 2007).

### 2.4 Covariates

Covariates extracted from clinical records included age at stroke onset (years), sex (male or female), stroke aetiology (ischaemic or haemorrhagic), stroke laterality (left or right hemisphere), time from stroke onset to RAGT initiation (days), baseline National Institutes of Health Stroke Scale (NIHSS) score, baseline Berg Balance Scale (BBS) score, number of RAGT sessions completed, exoskeleton platform (EksoNR or Indego), and rehabilitation setting (inpatient or outpatient at RAGT initiation). The modified Rankin Scale (mRS) at admission and the Charlson Comorbidity Index (CCI) were included as measures of global disability and comorbidity burden, respectively.

### 2.5 Statistical Analysis

#### 2.5.1 Joint Model Specification

A Bayesian shared-parameter joint model was specified consisting of two submodels linked through the subject-specific longitudinal trajectory. The longitudinal submodel for the FMA-LE trajectory of patient i at time t was:

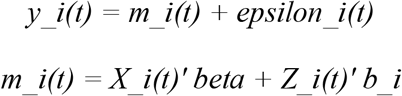

here y_i(t) denotes the observed FMA-LE score, m_i(t) is the true (unobserved) trajectory, X_i(t) contains fixed-effects covariates including a restricted cubic spline basis for time with three internal knots placed at weeks 4, 12, and 24, beta is the fixed-effects coefficient vector, Z_i(t) contains the random-effects design matrix (random intercept and random slope for time), b_i follows a multivariate normal distribution with mean zero and covariance matrix D, and epsilon_i(t) represents the measurement error assumed normally distributed with variance sigma squared.

The survival submodel for the hazard of achieving independent community ambulation was specified as a Weibull proportional hazards model:

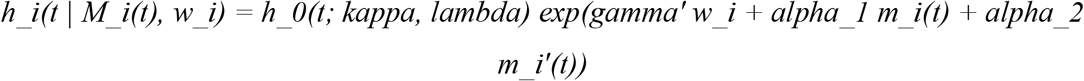

where h_0(t; kappa, lambda) is the Weibull baseline hazard with shape kappa and scale lambda, w_i is a vector of baseline covariates (age, sex, stroke aetiology, laterality, baseline NIHSS, baseline BBS, mRS, CCI, time to RAGT initiation, number of sessions, exoskeleton platform, and rehabilitation setting), gamma is the corresponding coefficient vector, m_i(t) is the current value of the subject-specific FMA-LE trajectory, m_i’(t) is the first derivative (slope) of the trajectory, and alpha_1 and alpha_2 are the association parameters linking the longitudinal and survival processes. The current-value association (alpha_1) quantifies how the current functional level relates to the instantaneous hazard, while the slope association (alpha_2) captures whether the rate of functional change provides additional predictive information beyond the current level.

#### 2.5.2 Prior Specification

Weakly informative priors were specified to allow the data to dominate inference while maintaining regularisation. Fixed-effects regression coefficients received normal priors with mean 0 and standard deviation 10. The Weibull shape parameter was assigned a gamma(2, 0.5) prior. The random-effects covariance matrix D was assigned an inverse-Wishart prior with degrees of freedom equal to the dimension plus one and a scaled identity matrix. The residual variance sigma squared received an inverse-gamma(0.01, 0.01) prior. Association parameters alpha_1 and alpha_2 received normal(0, 5) priors. A sensitivity analysis using more diffuse priors (standard deviations doubled) was conducted to assess prior influence.

#### 2.5.3 Estimation and Convergence

Posterior estimation was performed using Hamiltonian Monte Carlo (HMC) sampling implemented in Stan version 2.33 (Carpenter et al., 2017) called through the rstanarm package in R version 4.3.2 (Goodrich et al., 2024). Four chains of 5,000 iterations each were run with 2,500 warmup iterations, yielding 10,000 post-warmup draws for inference. Convergence was assessed using the Gelman-Rubin statistic (R-hat less than 1.05 for all parameters), effective sample sizes (minimum 400 per parameter), and visual inspection of trace plots. The leave-one-out information criterion (LOOIC) with Pareto smoothed importance sampling was used for model comparison between candidate longitudinal submodel specifications (linear, quadratic, and restricted cubic spline).

#### 2.5.4 Dynamic Predictions and Model Diagnostics

Dynamic predictions of conditional survival probabilities were computed for individual patients given their observed longitudinal data up to a landmark time s, providing updated milestone attainment probabilities as new FMA-LE measurements become available. Calibration was assessed using time-dependent area under the receiver operating characteristic curve (AUC) at landmark times of 4, 12, and 24 weeks, with 95% credible intervals obtained via posterior simulation. A five-fold cross-validation scheme was employed to evaluate out-of-sample prediction accuracy. Cox-Snell residuals from the survival submodel and standardised marginal residuals from the longitudinal submodel were examined for model adequacy.

#### 2.5.5 Sensitivity Analyses

Five sensitivity analyses were pre-specified: (a) alternative prior specifications with doubled standard deviations; (b) exclusion of patients who received fewer than 20 RAGT sessions; (c) a current-value-only joint model (omitting the slope association); (d) a piecewise-constant baseline hazard model as an alternative to the Weibull specification; and (e) stratification by exoskeleton platform (EksoNR versus Indego) to assess device-specific effects.

## 3. RESULTS

### 3.1 Study Population

Of 489 patients identified from institutional RAGT databases, 327 met all eligibility criteria and constituted the analytical sample. Reasons for exclusion included prior stroke (n = 62), fewer than two post-baseline FMA-LE assessments (n = 48), pre-existing neurological conditions (n = 31), and other contraindications (n = 21). The median age was 62.4 years (IQR 54.1 to 71.8), 190 (58.1%) were male, and 206 (63.0%) had ischaemic stroke aetiology. The median time from stroke onset to RAGT initiation was 48 days (IQR 28 to 82), and participants completed a median of 28 RAGT sessions (IQR 18 to 40). Baseline clinical characteristics are presented in Table 1. Patients who subsequently achieved independent community ambulation were younger, had lower NIHSS scores, higher baseline FMA-LE and BBS scores, and shorter intervals from stroke to RAGT initiation.

**Table 1.**
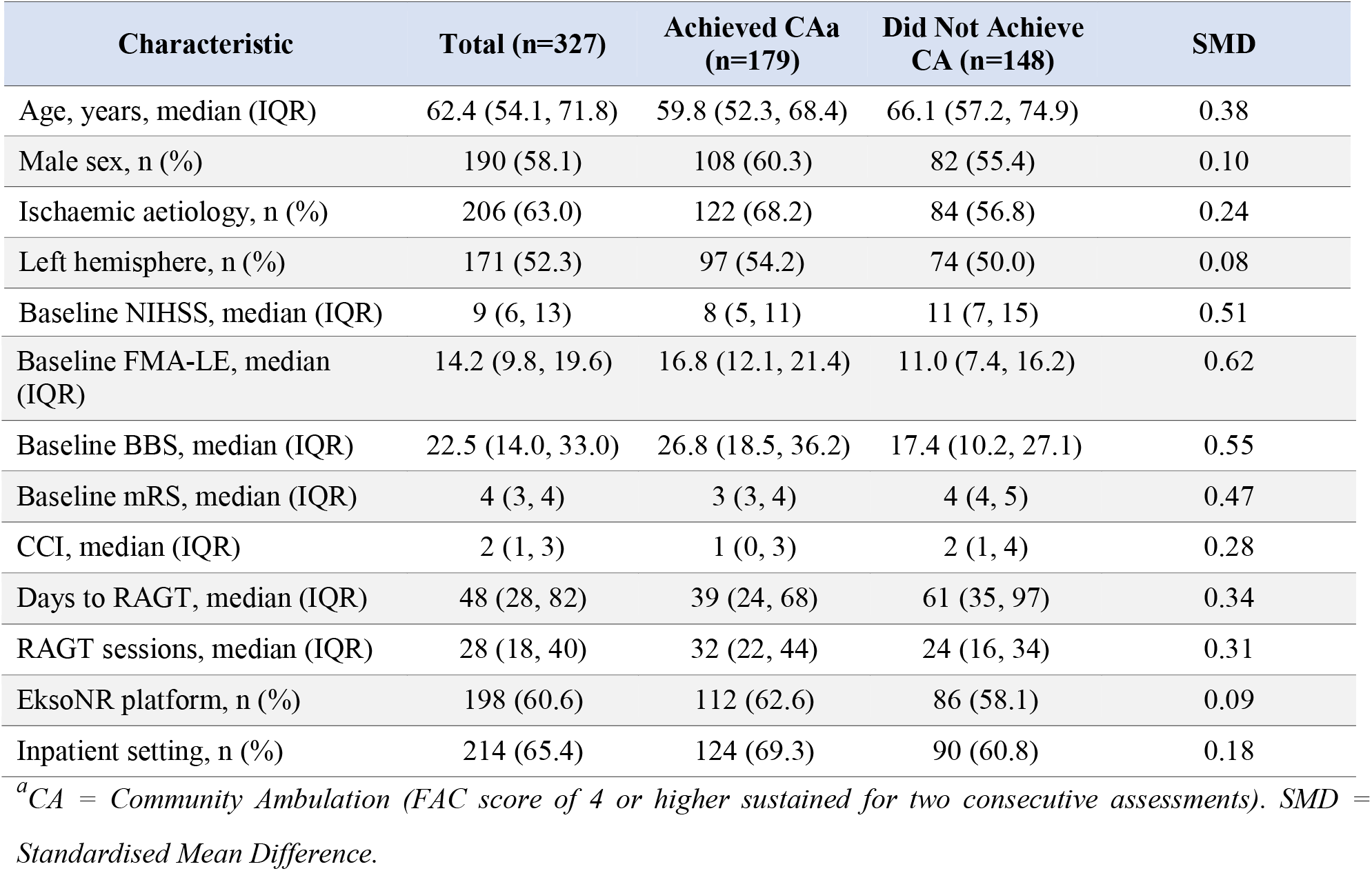
Baseline Characteristics of the Study Cohort Stratified by Community Ambulation Status.

| Characteristic | Total (n=327) | Achieved CAa (n=179) | Did Not Achieve CA (n=148) | SMD |
| --- | --- | --- | --- | --- |
| Age, years, median (IQR) | 62.4 (54.1, 71.8) | 59.8 (52.3, 68.4) | 66.1 (57.2, 74.9) | 0.38 |
| Male sex, n (%) | 190 (58.1) | 108 (60.3) | 82 (55.4) | 0.10 |
| Ischaemic aetiology, n (%) | 206 (63.0) | 122 (68.2) | 84 (56.8) | 0.24 |
| Left hemisphere, n (%) | 171 (52.3) | 97 (54.2) | 74 (50.0) | 0.08 |
| Baseline NIHSS, median (IQR) | 9 (6, 13) | 8 (5, 11) | 11 (7, 15) | 0.51 |
| Baseline FMA-LE, median (IQR) | 14.2 (9.8, 19.6) | 16.8 (12.1, 21.4) | 11.0 (7.4, 16.2) | 0.62 |
| Baseline BBS, median (IQR) | 22.5 (14.0, 33.0) | 26.8 (18.5, 36.2) | 17.4 (10.2, 27.1) | 0.55 |
| Baseline mRS, median (IQR) | 4 (3, 4) | 3 (3, 4) | 4 (4, 5) | 0.47 |
| CCI, median (IQR) | 2 (1, 3) | 1 (0, 3) | 2 (1, 4) | 0.28 |
| Days to RAGT, median (IQR) | 48 (28, 82) | 39 (24, 68) | 61 (35, 97) | 0.34 |
| RAGT sessions, median (IQR) | 28 (18, 40) | 32 (22, 44) | 24 (16, 34) | 0.31 |
| EksoNR platform, n (%) | 198 (60.6) | 112 (62.6) | 86 (58.1) | 0.09 |
| Inpatient setting, n (%) | 214 (65.4) | 124 (69.3) | 90 (60.8) | 0.18 |

<sup>a</sup>CA = Community Ambulation (FAC score of 4 or higher sustained for two consecutive assessments). SMD = Standardised Mean Difference.

### 3.2 Longitudinal FMA-LE Trajectory

A total of 1,634 FMA-LE observations were available across the 327 participants, with a median of 5 (IQR 4 to 6) assessments per patient. The overall completeness rate was 83.4%, with missing data primarily at the 52-week assessment (72.2% complete). Model comparison using LOOIC favoured the restricted cubic spline specification (LOOIC = 4,812.4) over the linear (LOOIC = 5,124.8) and quadratic (LOOIC = 4,967.2) alternatives.

The longitudinal submodel results are presented in Table 2. The restricted cubic spline coefficients indicated a distinctly nonlinear recovery trajectory characterised by rapid improvement during the first 12 weeks, with the steepest gains occurring between baseline and week 4 (spline basis 1 coefficient: 4.21, 95% CrI 3.28 to 5.16), continued but decelerating improvement from weeks 4 to 12 (coefficient: 3.64, 95% CrI 2.84 to 4.45), modest gains from weeks 12 to 24 (coefficient: 1.12, 95% CrI 0.39 to 1.85), and a near-plateau from weeks 24 to 52 (coefficient: 0.34, 95% CrI -0.23 to 0.91). The mean cumulative FMA-LE gain at 12 weeks was 8.7 points (95% CrI 7.2 to 10.3), and at 52 weeks was 9.8 points (95% CrI 7.9 to 11.8). Older age and higher baseline NIHSS were associated with lower trajectories, while haemorrhagic stroke aetiology was associated with a 2.14-point lower trajectory (95% CrI -3.28 to -1.01). Substantial between-patient variability was evident, with a random intercept standard deviation of 5.42 (95% CrI 4.78 to 6.12).

**Table 2.** Bayesian Longitudinal Submodel: Fixed and Random Effects for the FMA-LE Trajectory.

| Parameter | Mean | SD | 95% CrI | R-hat (ESS) |
| --- | --- | --- | --- | --- |
| Intercept | 14.38 | 0.62 | 13.17, 15.60 | 1.002 (8412) |
| Spline basis 1 (0 to 4 wk) | 4.21 | 0.48 | 3.28, 5.16 | 1.001 (7856) |
| Spline basis 2 (4 to 12 wk) | 3.64 | 0.41 | 2.84, 4.45 | 1.003 (7214) |
| Spline basis 3 (12 to 24 wk) | 1.12 | 0.37 | 0.39, 1.85 | 1.002 (8102) |
| Spline basis 4 (24 to 52 wk) | 0.34 | 0.29 | -0.23, 0.91 | 1.001 (8634) |
| Age (per 10 years) | -1.08 | 0.31 | -1.69, -0.48 | 1.002 (7918) |
| Male sex | 0.76 | 0.54 | -0.30, 1.82 | 1.001 (8256) |
| Haemorrhagic aetiology | -2.14 | 0.58 | -3.28, -1.01 | 1.003 (6892) |
| Baseline NIHSS (per point) | -0.38 | 0.09 | -0.56, -0.21 | 1.001 (8124) |
| Random intercept SD | 5.42 | 0.34 | 4.78, 6.12 | 1.004 (5834) |
| Random slope SD | 0.18 | 0.02 | 0.14, 0.22 | 1.003 (6124) |
| Residual SD (sigma) | 2.87 | 0.11 | 2.66, 3.09 | 1.001 (9012) |

*CrI = Credible Interval; SD = Standard Deviation; ESS = Effective Sample Size.*

### 3.3 Time to Independent Community Ambulation

During the 52-week follow-up period, 179 of 327 participants (54.7%) achieved independent community ambulation. The median time to milestone attainment was 18.4 weeks (95% CrI 15.8 to 21.2) among those who achieved it. The Kaplan-Meier estimated cumulative incidence was 22.3% at 12 weeks, 42.8% at 24 weeks, and 54.7% at 52 weeks. The Weibull shape parameter was 1.42 (95% CrI 1.22 to 1.65), indicating an increasing hazard over time consistent with the expectation that cumulative rehabilitation effects progressively enhance the probability of milestone attainment.

### 3.4 Joint Model: Association Parameters

The survival submodel results from the joint model are presented in Table 3. The current-value association parameter (alpha_1) was 0.084 (95% CrI 0.061 to 0.109), with a posterior probability exceeding 0.999 that the parameter is positive. This translates to a hazard ratio of 1.088 (95% CrI 1.063 to 1.115), indicating that each one-point increase in the subject-specific FMA-LE trajectory at any given time was associated with an 8.8% increase in the instantaneous hazard of achieving independent community ambulation. The trajectory slope association parameter (alpha_2) was 0.043 (95% CrI 0.012 to 0.078, posterior probability of being positive = 0.994), indicating that patients with steeper recovery gradients had a 4.4% additional increase in hazard per unit increase in slope, independent of their current functional level.

**Table 3.** Bayesian Survival Submodel: Association Parameters and Covariate Effects on Time to Community Ambulation.

| Parameter | Mean | SD | HR (95% CrI) | P(HR>1) | R-hat (ESS) |
| --- | --- | --- | --- | --- | --- |
| Association: Current value ($\alpha_1$) | 0.084 | 0.012 | 1.088 (1.063, 1.115) | >0.999 | 1.002 (7456) |
| Association: Slope ($\alpha_2$) | 0.043 | 0.017 | 1.044 (1.012, 1.081) | 0.994 | 1.003 (6812) |
| Age (per 10 years) | -0.211 | 0.074 | 0.81 (0.70, 0.94) | 0.003 | 1.001 (8124) |
| Male sex | 0.098 | 0.124 | 1.10 (0.87, 1.40) | 0.785 | 1.001 (8456) |
| Haemorrhagic aetiology | -0.386 | 0.162 | 0.68 (0.49, 0.93) | 0.010 | 1.002 (7234) |
| Left hemisphere | 0.054 | 0.118 | 1.06 (0.84, 1.33) | 0.675 | 1.001 (8312) |
| Baseline NIHSS (per point) | -0.062 | 0.016 | 0.94 (0.91, 0.97) | <0.001 | 1.002 (7856) |
| Baseline BBS (per 5 points) | 0.078 | 0.024 | 1.08 (1.03, 1.14) | 0.998 | 1.001 (8124) |
| Days to RAGT (per 30 days) | -0.094 | 0.038 | 0.91 (0.84, 0.98) | 0.008 | 1.002 (7654) |
| RAGT sessions (per 10) | 0.112 | 0.042 | 1.12 (1.03, 1.21) | 0.996 | 1.001 (7912) |
| EksoNR (vs Indego) | 0.068 | 0.128 | 1.07 (0.83, 1.37) | 0.700 | 1.001 (8234) |
| Weibull shape ($\kappa$) | 1.42 | 0.11 | (1.22, 1.65) | -- | 1.002 (7456) |

HR = Hazard Ratio; CrI = Credible Interval; P(HR>1) = Posterior probability that HR exceeds 1; ESS = Effective Sample Size.

Among the baseline covariates in the survival submodel, haemorrhagic stroke aetiology (HR = 0.68, 95% CrI 0.49 to 0.93), older age (HR per decade = 0.81, 95% CrI 0.70 to 0.94), and higher baseline NIHSS (HR per point = 0.94, 95% CrI 0.91 to 0.97) were associated with lower hazards of achieving the ambulation milestone. Longer time from stroke onset to RAGT initiation was also associated with reduced hazard (HR per 30 days = 0.91, 95% CrI 0.84 to 0.98), supporting early initiation of RAGT. Higher baseline Berg Balance Scale scores (HR per 5 points = 1.08, 95% CrI 1.03 to 1.14) and greater number of RAGT sessions (HR per 10 sessions = 1.12, 95% CrI 1.03 to 1.21) were associated with higher hazards of milestone attainment. No statistically credible differences were observed between exoskeleton platforms (HR = 1.07, 95% CrI 0.83 to 1.37) or sex (HR = 1.10, 95% CrI 0.87 to 1.40).

### 3.5 Dynamic Predictions

The joint model demonstrated good discriminatory performance for dynamic predictions, with AUC values improving as more longitudinal data became available (Table 4). At the 4-week landmark predicting 12-week outcomes, the time-dependent AUC was 0.78 (95% CrI 0.73 to 0.83). This improved to 0.83 (95% CrI 0.78 to 0.88) at the 12-week landmark for 24-week predictions and reached 0.87 (95% CrI 0.82 to 0.91) at the 24-week landmark for 52-week predictions. Five-fold cross-validated Brier scores correspondingly decreased with later landmark times, ranging from 0.182 (4-week landmark) to 0.128 (24-week landmark), indicating progressively better calibration as the longitudinal history accumulated.

**Table 4.** Dynamic Prediction Performance of the Bayesian Joint Model at Various Landmark Times.

| Landmark Time | Prediction Horizon | AUC (95% CrI) | Brier Score (95% CrI) |
| --- | --- | --- | --- |
| 4 weeks | 12 weeks | 0.78 (0.73, 0.83) | 0.182 (0.158, 0.208) |
| 4 weeks | 24 weeks | 0.74 (0.68, 0.79) | 0.201 (0.174, 0.229) |
| 12 weeks | 24 weeks | 0.83 (0.78, 0.88) | 0.152 (0.128, 0.178) |
| 12 weeks | 52 weeks | 0.81 (0.76, 0.86) | 0.167 (0.142, 0.194) |
| 24 weeks | 52 weeks | 0.87 (0.82, 0.91) | 0.128 (0.104, 0.154) |

*AUC = Area Under the Receiver Operating Characteristic Curve; CrI = Credible Interval.*

### 3.6 Sensitivity Analyses

Results from the five pre-specified sensitivity analyses are summarised in Table 5. The association parameters remained stable across all specifications. Using diffuse priors (doubled standard deviations) yielded virtually identical estimates (alpha_1 = 0.081, 95% CrI 0.057 to 0.106), confirming that the posterior was data-driven. Restricting the sample to patients with 20 or more RAGT sessions (n = 248) slightly strengthened the associations (alpha_1 = 0.091, alpha_2 = 0.048), consistent with greater treatment exposure amplifying the trajectory-outcome relationship. The current-value-only model (omitting alpha_2) resulted in a worse LOOIC (4,834.6 versus 4,812.4), supporting the incremental predictive value of the slope parameter. The piecewise-constant baseline hazard specification produced consistent estimates. Stratification by exoskeleton platform showed consistent associations for both EksoNR and Indego subgroups, although the Indego subgroup exhibited wider credible intervals due to smaller sample size.

**Table 5.**
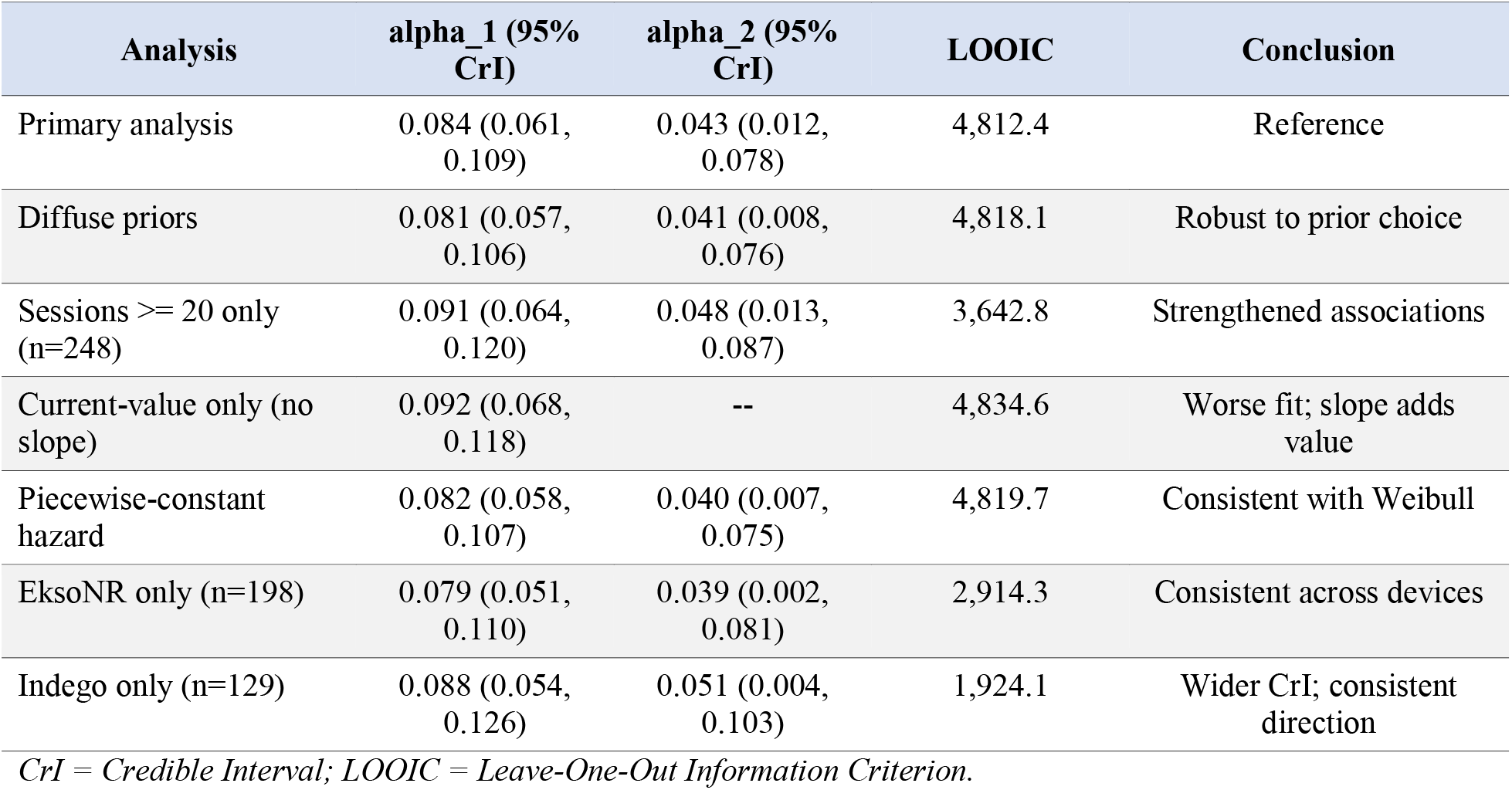
Sensitivity Analyses: Association Parameter Estimates Under Alternative Model Specifications.

## 4. DISCUSSION

This study applied a Bayesian joint longitudinal-survival model to characterise the association between functional recovery trajectories and time to independent community ambulation following robotic exoskeleton-assisted gait training in a multi-centre Canadian stroke rehabilitation cohort. The key findings were threefold. First, the FMA-LE recovery trajectory following RAGT was distinctly nonlinear, with the majority of gains occurring within the initial 12 weeks. Second, both the current functional level and the rate of functional improvement were independently and positively associated with the hazard of achieving community ambulation, with the current-value association demonstrating a stronger effect. Third, the joint model exhibited good dynamic prediction performance that improved progressively as longitudinal data accumulated, suggesting clinical utility for real-time prognostication.

The nonlinear recovery trajectory observed in this study is consistent with the broader stroke rehabilitation literature, which has consistently demonstrated a logarithmic or exponential recovery pattern with the greatest gains occurring in the first 3 months post-stroke (Kwakkel et al., 2004; Prabhakaran et al., 2008). However, few studies have examined this pattern specifically in the context of RAGT. The mean 12-week FMA-LE gain of 8.7 points in our cohort exceeds the minimal clinically important difference of 6 points reported by Page et al. (2012) and is comparable to or slightly higher than gains reported in randomised controlled trials of RAGT (Molteni et al., 2021). This aligns with findings from Shankar et al. (2025), who demonstrated favourable cost-effectiveness of robotic exoskeleton therapy for stroke rehabilitation, suggesting that the functional gains achieved with RAGT translate into both clinically meaningful and economically justifiable outcomes.

The significant association between the subject-specific FMA-LE trajectory and the hazard of community ambulation represents a novel contribution to the rehabilitation literature. The current-value association (HR = 1.088 per FMA-LE point) indicates that functional level at any given time is a strong determinant of milestone attainment, while the additional slope association (HR = 1.044 per unit slope) suggests that the velocity of recovery carries independent prognostic information. This has direct clinical implications: two patients at the same FMA-LE score may have substantially different probabilities of achieving community ambulation depending on whether their trajectory is ascending, plateauing, or declining. Clinicians may use this information to identify patients who are on a favourable trajectory and may benefit from sustained RAGT intensity, as well as patients whose trajectories are plateauing and may benefit from alternative or augmented interventions.

The finding that haemorrhagic stroke aetiology was associated with both lower longitudinal trajectories and reduced hazard of community ambulation is consistent with prior reports documenting greater initial severity and slower recovery in haemorrhagic compared with ischaemic stroke (Paolucci et al., 2003). Similarly, the adverse associations of older age and higher baseline NIHSS with milestone attainment are well established in the stroke prognostication literature (Veerbeek et al., 2011). The protective association of earlier RAGT initiation (HR per 30-day delay = 0.91) supports contemporary practice guidelines recommending early and intensive rehabilitation (Winstein et al., 2016), though the retrospective design precludes definitive causal claims.

The dynamic prediction framework demonstrated in this study has potential translational value for personalised rehabilitation planning. The ability to update a patient’s predicted probability of achieving community ambulation as new FMA-LE assessments become available could support shared decision-making regarding treatment intensity, duration, and discharge planning. The AUC of 0.87 at the 24-week landmark for 52-week predictions suggests that the joint model captures substantial prognostic information. This framework is particularly relevant in contexts where RAGT resources are constrained and optimal allocation requires evidence-based prognostication, a consideration highlighted by the economic evaluation of Shankar et al. (2025), who demonstrated that cost-effectiveness was most favourable when robotic exoskeleton therapy was targeted to patients with the highest likelihood of meaningful functional gains.

### 4.1 Strengths and Limitations

This study has several strengths. The Bayesian joint modelling framework simultaneously addresses informative dropout and the endogenous nature of time-varying functional measures, producing less biased estimates than separate longitudinal and survival analyses. The multi-centre design across four Canadian provinces enhances generalisability within the Canadian healthcare context. The flexible nonlinear specification of the longitudinal trajectory avoids the potentially misleading assumption of linear recovery. The comprehensive sensitivity analyses demonstrate robustness of the findings to alternative model specifications, prior choices, and subgroup restrictions.

Several limitations warrant consideration. First, the retrospective design introduces potential for confounding and selection bias, as patients receiving RAGT may differ systematically from those receiving conventional therapy. Second, the FAC-based definition of community ambulation, while clinically meaningful, does not capture aspects of ambulation quality such as endurance, symmetry, and safety. Third, the 52-week follow-up may not capture late recoveries, and the declining assessment completeness at later timepoints could introduce informative missingness despite the joint model’s partial accommodation of this issue. Fourth, the sample size, while adequate for the primary joint model, limited statistical power in the platform-stratified sensitivity analysis. Fifth, the analysis did not account for concomitant therapies (e.g., conventional physiotherapy, occupational therapy) that participants likely received alongside RAGT. Finally, the study was conducted within the Canadian public healthcare system, and generalisability to other healthcare contexts with different rehabilitation service delivery models should be considered carefully.

### 4.2 Implications for Practice and Research

The findings support several clinical recommendations. First, rehabilitation clinicians should consider both the current functional level and the rate of change when prognosticating outcomes for stroke patients undergoing RAGT. A patient demonstrating rapid early improvement may warrant continued intensive RAGT, whereas a patient whose trajectory has plateaued may benefit from reassessment and potential modification of the rehabilitation programme. Second, the dynamic prediction framework could be operationalised as a clinical decision support tool, providing real-time updated probabilities of milestone attainment to support shared decision-making. Third, the finding that earlier RAGT initiation was associated with better outcomes reinforces the importance of timely access to robotic rehabilitation technologies.

Future research should prioritise prospective validation of the joint model in independent cohorts, incorporation of additional biomarkers (e.g., neuroimaging, neurophysiological measures) as longitudinal predictors, and extension to competing risks frameworks that account for multiple rehabilitation milestones simultaneously. The integration of joint modelling with health economic evaluations, building on the cost-effectiveness framework established by Shankar et al. (2025), would provide a comprehensive evidence base for personalised resource allocation in robotic stroke rehabilitation.

## 5. CONCLUSIONS

This multi-centre Canadian cohort study demonstrated that a Bayesian joint longitudinal-survival model effectively characterises the interdependence between functional recovery trajectories and time to independent community ambulation following robotic exoskeleton-assisted gait training for stroke. Both the current functional level and the velocity of recovery independently predicted milestone attainment, supporting personalised rehabilitation planning based on the evolving trajectory. The dynamic prediction framework showed progressively improving discrimination as longitudinal data accumulated, suggesting clinical utility for real-time prognostication. These findings contribute to the growing evidence base for robotic exoskeleton interventions in stroke rehabilitation and provide a methodological template for future studies examining trajectory-outcome associations in rehabilitation medicine.

## Data Availability

All data produced in the present work are contained in the manuscript

## DECLARATIONS

### Ethics Approval

This study was approved by the Research Ethics Board at each participating institution (lead REB: University Health Network, REB No. 2023-0412).

### Consent to Participate

A waiver of individual consent was granted given the retrospective design using de-identified clinical data.

### Data Availability

The data underlying this study contain identifiable clinical information and cannot be shared publicly. Requests for de-identified aggregate data may be directed to the corresponding author.

### Author Contributions

CMT conceived the study, led data analysis, and drafted the manuscript. AJB contributed to the statistical methodology and Bayesian modelling. PRD contributed to outcome measure selection and clinical interpretation. JLM oversaw data collection at the Toronto site and contributed to clinical interpretation. SKW contributed to the study design and health economic considerations. MAG contributed to data extraction and quality assurance. All authors reviewed and approved the final manuscript.

